# Knowledge and attitude towards breast cancer screening among female students at the University of Zambia: a cross-sectional study

**DOI:** 10.1101/2025.11.19.25340605

**Authors:** Mwansa Ketty Lubeya, Clemency Hamvumba, Luwi Mercy Mwangu, Innocent Maposa, Ellah Zingani, Moses Mukosha

## Abstract

**Introduction:** Breast cancer is a significant public health concern worldwide. It is the second leading cause of cancer-related death in Zambia. A major contributor to the high morbidity and mortality is late-stage diagnosis due to a lack of knowledge of the risk factors, early signs and symptoms, and breast cancer screening methods. This study aimed to assess the level of Knowledge and Attitude towards breast cancer screening methods among female students at the University of Zambia and determine the factors associated with the knowledge and attitude towards breast cancer screening

**Methods:** This cross-sectional study was conducted 1^st^ August and 30^th^ September 2024. A previously validated structured questionnaire was used to collect data. The data from the online questionnaire was automatically entered into Microsoft Excel. The data were coded and analysed using Stata. Logistic regression determined factors influencing knowledge and attitude, adjusting for other variables.

**Results:** Of the 389 respondents, 39 (10.0%) were not aware of breast cancer. Of those who were aware, 132 (38.8%) had good knowledge, and 189 (55.6%) had a good attitude. Compared to first-year students, fifth-year students (adjusted odds ratio [aOR]= 3.74, 95% CI: 1.34, 10.48) and sixth-year or above students (aOR=5.45, 95% CI: 1.65, 18.04) were more likely to have good knowledge.

On the other hand, students in non-health-related training programs (aOR=0.22, 95% CI: 0.13, 0.38) were less likely to have good knowledge than students from health-related training programs. Similarly, students in non-health-related training programs (aOR=0.49, 95% CI: 0.30, 0.79) were less likely to have good attitudes than students from health-related training programs.

**Conclusion:** This study reports poor knowledge and moderate attitudes among university students. Since good knowledge and attitude are strongly linked to successful breast cancer awareness programs, future implementation studies should target female students from non-health-related training programs for increased breast cancer screening uptake.

## Introduction

Breast cancer is a prevalent global health problem and a major cause of death in women [1]. This kind of cancer can range from non-invasive to metastatic carcinoma, with its origins in the breast tissue [2]. According to the World Health Organisation, almost 2.3 million cases of breast cancer were diagnosed globally in 2020 [2, 3]. Low and middle-income countries share a disproportionate burden, accounting for over 74% of the global burden of disability-adjusted life years lost due to breast cancer [4].

In Zambia, breast cancer accounts for 8% of all cancer patients seen at the Cancer Diseases Hospital, making it the second most frequent malignancy among women after cervical cancer [5]. Lifestyle changes and increased urbanisation are potentially contributing factors to the rising incidence of breast cancer in developing economies [6].

For effective breast cancer screening and early diagnosis, sufficient knowledge and awareness among healthcare workers and the general public are of utmost importance in reducing breast cancer cases [7]. Reducing breast cancer-related morbidity and mortality requires adequate knowledge of the disease’s signs and symptoms as well as early detection through self-breast examination, clinical breast examination, or Mammogram and other screening techniques [8].

Screening has been shown to reduce cancer death rates by 25 – 30 % [9]. However, in most developing countries, there is a lack of early detection programs and suitable facilities for diagnosis and treatment [10]. Due to delays in symptom recognition and diagnosis, two-thirds of female patients with breast cancer present at the hospital with late-stage disease, which raises the mortality rate [11, 12].

The Ministry of Health in Zambia has established cancer awareness and prevention programs, including self-breast examination, clinical breast examination, breast ultrasound, and guided breast biopsy, in certain parts of the country [13]. Breast self-examination is the least expensive early detection method, as it does not require advanced knowledge or physician intervention, and has been encouraged as part of awareness programs in Zambia [13].

Despite these preventive efforts by the Ministry of Health, there are still existing gaps, as seen by the high prevalence of breast cancer in Zambia of 19.9 cases per 100,000 women, and the mortality rate at 8.5 deaths per 100,000 women. [14]. To address this gap, this study aimed to determine predictors and assess the level of knowledge and attitude towards breast cancer screening and self-breast examination among female students at the University of Zambia.

Studies have shown that having a good attitude and knowledge about breast cancer screening and self-breast examination reduces the incidence and mortality rate of breast cancer [14]. Despite this, the attitude and knowledge levels in Zambia remain critically low. Thus, this study assessed the level of knowledge and attitude towards breast cancer screening and self-breast examination among female students at the University of Zambia.

## Methods

### Study design, setting and population

This was a cross-sectional study of female students conducted between 1^st^ August and 30^th^ September 2024. The study targeted the University of Zambia female students. This study population represents mostly youthful women from different parts of the country, including both rural and urban settings. This study included students aged 18 and above who were willing to consent to be part of the study. The study excluded students who reported not being aware of breast cancer.

### Sample Size Determination

The sample size was determined using the Cochran formula for sample size estimation. We assumed the prevalence of good knowledge of breast cancer of 49.7%, based on a similar study by Zimba et al [15], a sampling error of 5%, and 95% confidence intervals. The final sample size was 389 after adjusting for non-response

### Data Collection Tool

A previously validated questionnaire was adapted from a similar study done in Zambia [15] at the household level and the Kingdom of Saudi Arabia [7]. The resulting questionnaire was pilot-tested after content and face validation from experts based at the Cancer Diseases Hospital and the University of Zambia. After some minor modifications, the final questionnaire had three sections.

Section A, Socio-demographic information; age, year of study (year 1 to seven or postgraduate), Residence (rural or urban), Marital status (married or unmarried), Religion and program of study. Section B, consisted of 11 Knowledge questions on breast cancer screening and self-examination.

Section C, consisted of eight questions on attitude towards breast cancer screening. Knowledge and Attitude were conceptualised as defining breast cancer, identifying screening methods, their importance, frequency, eligibility for each method, and practices of self-breast exams. The continuous scores for knowledge and attitude were categorised using Bloom’s taxonomy of educational objectives into good (scores ≥60%) and poor (scores <60%).

Information about the study was shared through various students’ WhatsApp groups and associations. The questionnaire was administered to the respondents electronically via the Google forms@ platform. In addition, respondents were sent emails with an information sheet, consent form and the questionnaire. Written informed consent was obtained from all study participants. Overall, 700 questionnaires were sent out, of which 389 (55.6%) were returned. Follow-up messages were sent every 2 weeks, three times to ensure a high response rate.

### Statistical analysis

Data collected from respondents was imported into Excel, cleaned, coded and analysed thereafter using Stata/BE version 18 (Stata Corporation, College Station, TX, USA). The data were described using frequencies with percentages for categorical variables and means with standard deviations for continuous variables. Pearson Chi-square test or Fisher’s exact test, as appropriate, were used to assess the association between Knowledge or Attitude and independent variables.

Variables to include in the multivariable models were selected based on the researcher’s experience and those with significance of less than 20% from the univariate model. Two separate logistic regression models were fitted to determine factors associated with knowledge and attitude towards breast cancer screening, adjusting for other variables. The statistical significance was set at alpha less than 0.05, and all tests were two-sided.

### Ethical Consideration

Ethical approval was sought from the University of Zambia Health Sciences Research Ethics Committee (UNZAHSREC, ref 20231270208). Additional permission was granted by the Zambia National Health Research Authority (NHRA) and the University of Zambia management. The purpose of the research was clearly explained to the participants using the same platforms to share the link to the questionnaire. Participation was voluntary, and participants were informed of their right to withdraw at any time without fear of any repercussions.

Confidentiality was enforced to the extent legally acceptable. There was no direct benefit of participating in this study. However, findings could inform policymakers initiating and implementing breast cancer awareness programs in Zambia and similar settings.

## Results

Table 1 shows the characteristics of study participants. We enrolled 389 participants; 49 (12.8%) were unaware of breast cancer and were dropped from further analysis. The majority, 268 (78.8%), were aged between 20 and 29 years, and 327 (96.2%) were unmarried. Slightly above half, 171 (50.3%), were in health-related training programs, and about one in five, 77 (22.7%), were in their first year of study.

**Table 1.**
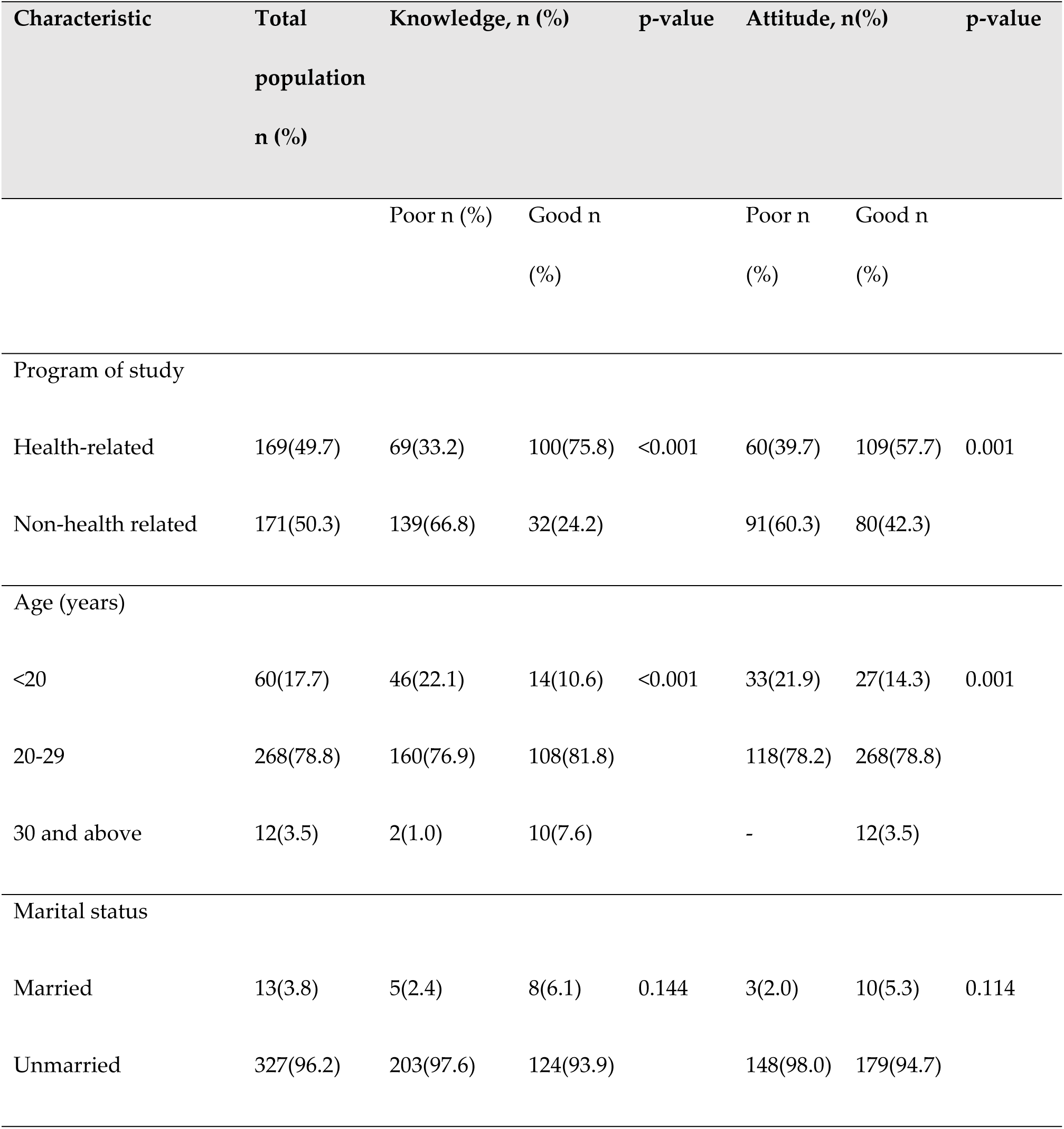
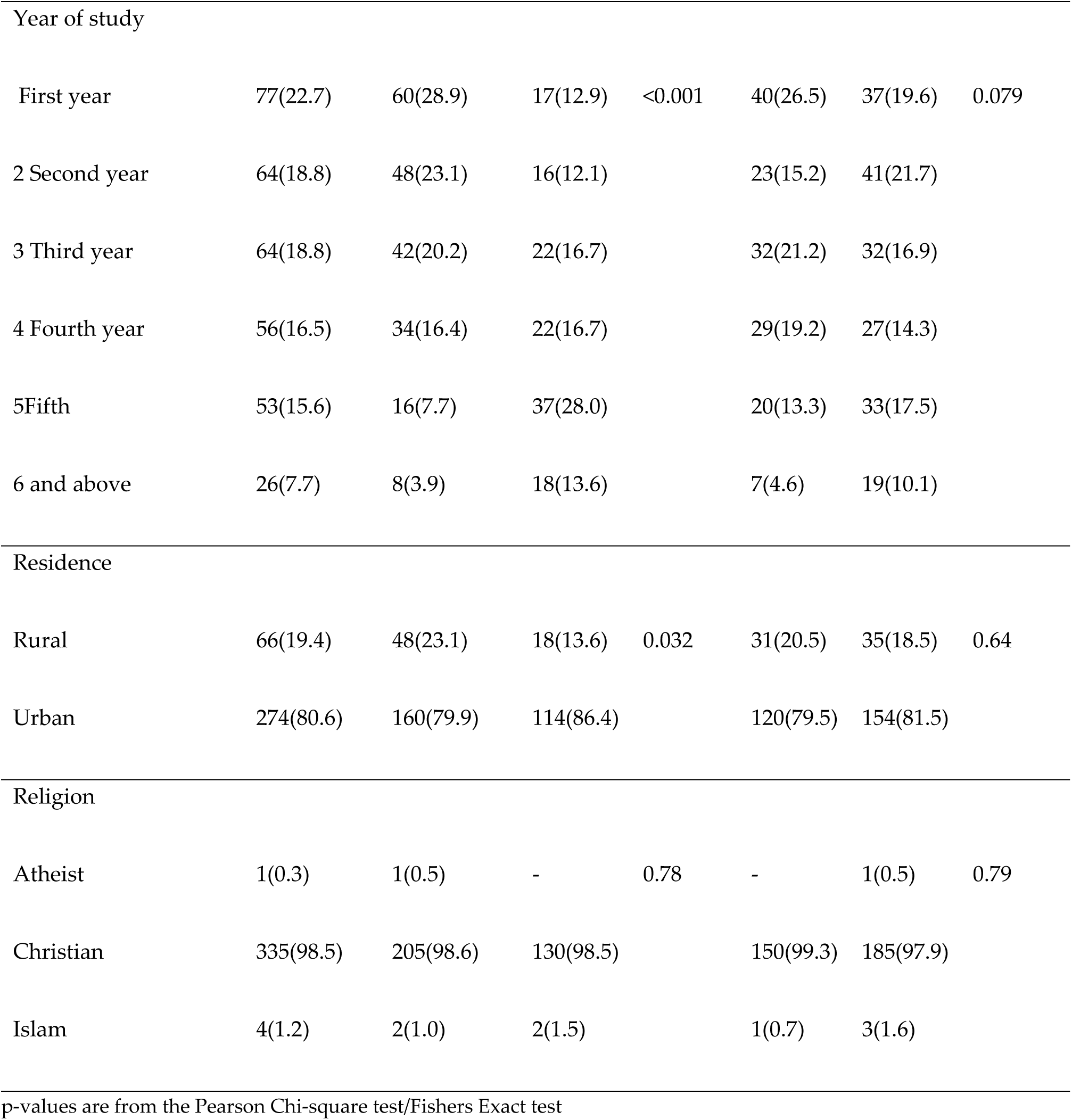
Characteristics of respondents by knowledge and attitude towards breast cancer at the University of Zambia (N=340)

Additionally, 274 (80.6%) were from urban areas. There was evidence of an independent association between the knowledge of breast cancer and age, program of study, year of study and residence. Furthermore, there was an association between attitude, age and program of study.

Among students who had poor knowledge, 66.8% were from non-health-related programs of study, 76.6% were aged between 20-29, 97.6% were unmarried, and 79.9% were from urban areas. Additionally, as the year of study increases, the proportion of people with poor knowledge decreases.

### Knowledge of Breast Cancer Screening

Overall, from the 340 respondents who were aware of breast cancer 132 (38.8%) had good knowledge of breast cancer screening and breast self-examination. Most of the respondents, 306 (90.0%) answered correctly what breast cancer screening is, 113 (33.2%) knew who can get breast cancer, 137 (40.3%) thought breast cancer can be prevented, and 317 (93.2%) agreed that early breast cancer screening can improve treatment success rate. About one in five respondents 61 (17.9%), correctly reported how often one should do breast self-examination, and half 170 (50.0%), correctly reported at what age self-breast examination should start. Slightly below half of respondents, 145(42.6%) agreed that Mammography is useful for early detection of breast cancer, 18(5.3%) correctly reported the age at which women should start regular Mammograms, and 289(84.1%) reported that clinical breast exam by a healthcare provider is necessary even if you regularly perform breast self-examinations (Table 2).

**Table 2.0:**
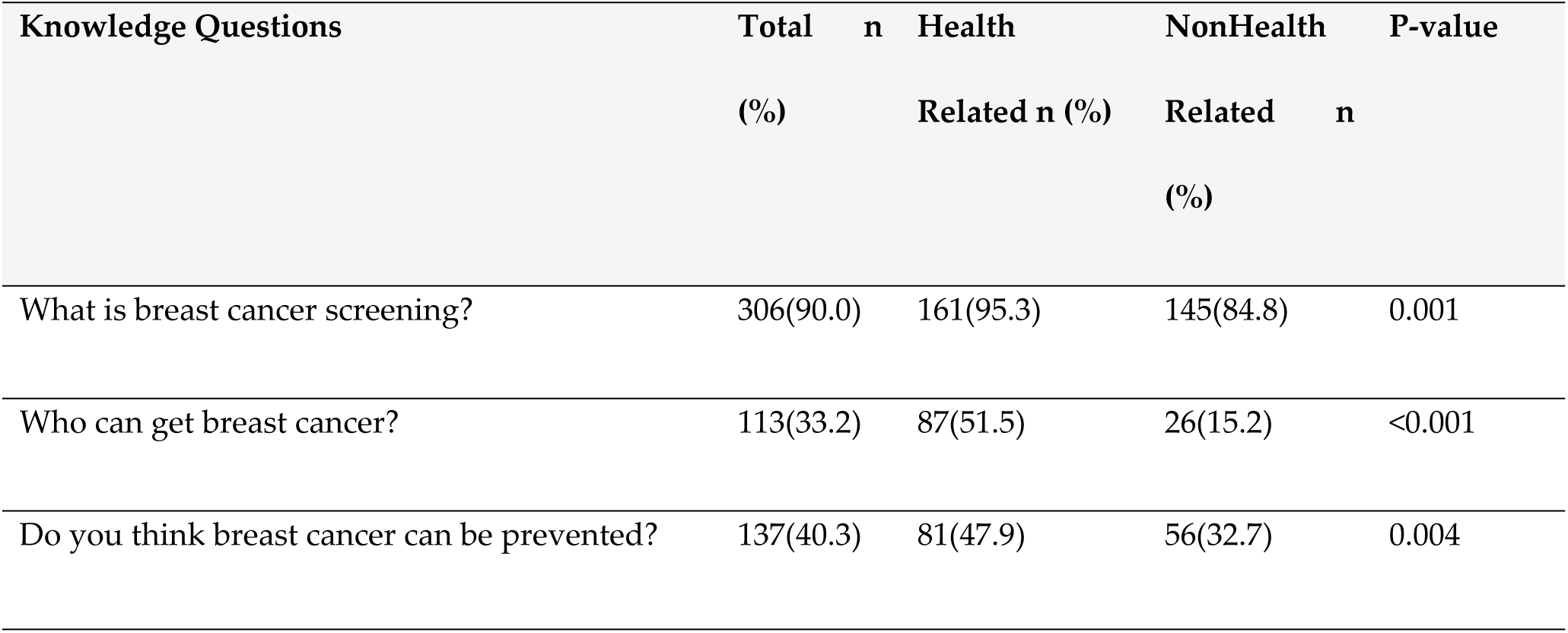
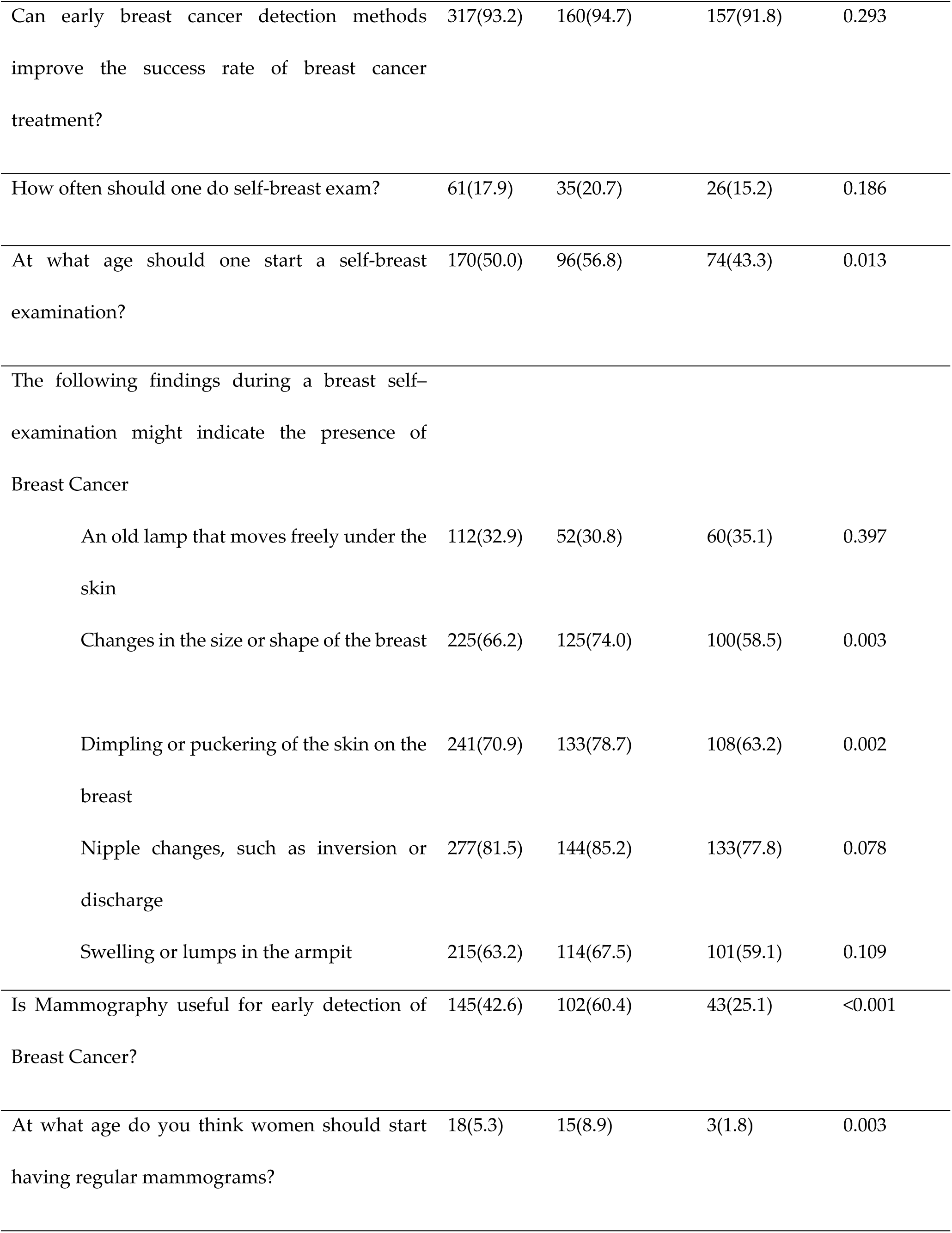
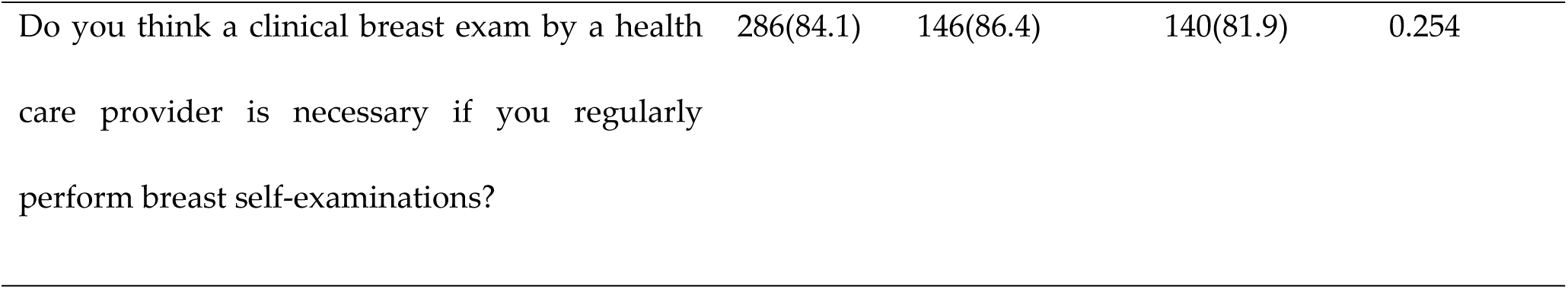
Proportion of Correct Responses to Knowledge Questions.

Overall, participants from health-related training programs reported the highest proportion of participants with good knowledge of breast cancer screening: 100/169 (59.2%) compared to those from non-health-related training programs: 32/171 (18.7%).

### Attitude towards breast cancer screening

The overall attitude towards breast cancer screening was 189 (55.6%). Slightly below two in five 126, 37.1%) reported doing a breast examination, 307(90.3%) reported that they would see a doctor upon detection of breast abnormality on self-breast examination, and 8(2.4%) have had a clinical breast examination conducted by a healthcare provider regularly.

Additionally, 135(39.7%) of respondents reported being very concerned about developing breast cancer, 59(17.4%) indicated a strong intention to undergo breast cancer screening, 294(86.5%) felt that regular breast check-ups are necessary even if you don’t have any breast problems, 317(93.2%) believed that breast cancer screening can save lives, and 10(2.9%) reported that they have undergone mammography before (Table 3).

**Table 3.** Proportion of Correct Responses to Attitude Questions.

| Attitude Questions | Total<br>n(%) | Health<br>Related n(%) | Non Health<br>Related n(%) | P-value |
| --- | --- | --- | --- | --- |
| Do you do a Self-Breast Examination? | 126(37.1) | 86(50.9) | 40(23.4) | <0.001 |
| What action would you take upon detection of breast abnormality on self-breast examination? | 307(90.3) | 157(92.9) | 150(87.7) | 0.107 |
| Have you ever had a clinical breast exam conducted by a healthcare provider? | 8(2.4) | 5(3.0) | 3(1.8) | 0.409 |
| How concerned or worried are you about the possibility of developing breast cancer? | 135(39.7) | 65(38.5) | 70(40.9) | 0.353 |
| How likely are you to have a screening clinical breast examination, ultrasound or mammography in the next 12 months? | 59(17.4) | 34(20.7) | 24(14.0) | 0.383 |
| Do you believe that regular breast check-ups are necessary even if you do not have any breast problems? | 294(86.5) | 154(91.1) | 140(81.9) | 0.013 |
| Do you believe breast cancer screening can save lives? | 317(93.2) | 158(93.5) | 159(93.0) | 0.852 |
| Have you ever undergone a mammography? | 10(2.9) | 5(3.0) | 5(2.9) | 1.000 |

Overall, participants from health-related training programs reported the highest proportion of participants with good Attitudes towards breast cancer screening: 109/169 (64.5%) compared to those from non-health-related training programs: 80/171 (46.8%).

### Predictors of breast cancer knowledge and attitude among students

In multivariable analysis, factors associated with breast cancer good knowledge were year of study and program of study. On the other hand, factors associated with a good attitude towards breast cancer screening and self-examination were the program of study. Compared to first-year students, fifth-year students (adjusted odds ratio [aOR]= 3.74, 95% CI: 1.34, 10.48) and sixth-year or above students (aOR=5.45, 95% CI: 1.65, 18.04) were more likely to have good knowledge.

On the other hand, students in non-health-related training programs (aOR=0.22, 95% CI: 0.13, 0.38) were 78% less likely to have good knowledge than students from health-related training programs. Similarly, students in non-health-related training programs (aOR=0.49, 95% CI: 0.30, 0.79) were 51% less likely to have good attitudes than students from health-related training programs (Table 4).

**Table 4.** Factors associated with good knowledge and attitude towards breast cancer.

| Variable | Good Knowledge, n=132(38.8%) |  | Good Attitude, n=189(55.64%) |  |
| --- | --- | --- | --- | --- |
|  | aOR (95% CI) | p-value | aOR (95% CI) | p-value |
| Program of study |  |  |  |  |
| Health-related | Ref |  | Ref |  |
| Non-health related | 0.22(0.13, 0.38) | <0.001 | 0.49(0.30, 0.79) | 0.003 |
| Age (years) |  |  |  |  |
| <20 | Ref |  |  |  |
| 20-29 | 0.91(0.35, 2.32) | 0.838 |  |  |
| 30 and above | 5.05(0.77, 33.0) | 0.091 |  |  |
| Marital status |  |  |  |  |
| Married |  |  | Ref |  |
| Unmarried |  |  | 0.43(0.11, 1.66) | 0.220 |
| Year of Study |  |  |  |  |
| 1 | Ref |  | Ref |  |
| 2 | 1.10(0.42, 2.84) | 0.847 | 1.81(0.91, 3.60) | 0.093 |
| 3 | 1.28(0.48, 3.40) | 0.624 | 0.88(0.45, 1.76) | 0.725 |
| 4 | 1.80(0.66, 4.93) | 0.250 | 0.84(0.41, 1.70) | 0.625 |
| 5 | 3.74(1.34, 10.48) | 0.012 | 1.10(0.51, 2.41) | 0.803 |
| 6 and above | 5.45(1.65, 18.04) | 0.005 | 1.95(0.70, 5.42) | 0.199 |
| Residence |  |  |  |  |
| Rural | Ref |  |  |  |
| Urban | 1.53(0.79, 2.95) | 0.209 |  |  |

## Discussion

The distribution of knowledge and attitude towards breast cancer screening and self-breast examination across different variables, as well as the factors associated with them, were analysed among female students at the University of Zambia. Overall, about two in five of the respondents had good knowledge, with slightly below three in five reporting a good attitude. The knowledge and attitude were higher among respondents from the health-related training programs.

In adjusted analysis, the odds of having good knowledge were lower among respondents from non-health-related than health-related programs and for fifth and sixth-year students than first-year students. Additionally, the odds of a good attitude were higher for participants in health-related training programs than those in non-health-related training programs.

The findings reveal that poor knowledge of breast cancer was most prevalent among specific subgroups. A majority (66.8%) of students with poor knowledge were enrolled in non-health-related programs, highlighting the potential lack of exposure to health education in those fields. Additionally, poor knowledge was more common among students aged 20–29 (76.6%), unmarried (97.6%), and those from urban areas (79.9%). Interestingly, despite better access to information in urban settings, urban residency did not appear to translate into higher awareness.

These patterns suggest that targeted awareness campaigns may be needed for younger, unmarried, urban-dwelling students outside health-related disciplines. Notably, there was a clear inverse relationship between year of study and poor knowledge: as students progressed in their academic years, the proportion with poor knowledge declined. This trend indicates that academic exposure over time may contribute to increased awareness, underscoring the value of integrating breast cancer education earlier in university curricula, especially for students outside the health sciences.

The proportion of respondents with good knowledge of breast cancer screening in this study was in line with similar studies [16, 17]. For instance, Ibnawadh and colleagues reported that university students showed a lack of knowledge in breast cancer self-examination, even though their attitude towards it was positive [18]. However, these findings were contrary of Olufemi and colleagues, who found that 62.2% of University students had good knowledge of breast cancer screening [19].

In developed countries, the proportions have been varied but consistently higher than in developing countries, like the current study setting [20]. The reason could be in different levels of education or sensitisation programs among students from different regions of the world. For instance, in the United Kingdom, breast cancer has drastically reduced due to the robust sensitisation programs introduced in 2015 [21].

The overall percentage of respondents with good attitudes was similar to the extant literature [22]. A study by Pal and colleagues found that about 71.1% had good attitudes towards breast cancer screening [23]. Similarly, Olufemi and colleagues reported good attitudes toward breast cancer screening among university students [19]. Also, Yusuf and colleagues reported that more than 50% of the participants had good Attitudes towards breast cancer screening [24].

However, some studies have reported a poor attitude towards breast cancer screening. Kinteh and colleagues found that 18% of the respondents had good attitudes towards breast cancer screening [25]. Based on the traditional education theory, knowledge levels directly affect attitude levels, which, in turn, affect practice. The difference in attitude among the reported studies could be explained by the knowledge levels. For example, another study [25] knowledge levels were below average, which could explain the moderate attitude levels observed, similar to observations by Pal and colleagues [23].

In this study, a program of study was found to predict the odds of having knowledge and attitude towards breast cancer screening and self-examination. These findings were consistent with a study conducted among female students at the University of Sharjah, where they noted an association between knowledge of breast cancer screening and the program of study [26].

According to the study, participants from health-related training programs were more knowledgeable about breast cancer screening than those from non-health-related programs [26]. These findings were corroborated by a study conducted in Pakistan by Jawaria and colleagues among female medical and non-medical students, which reported that medical students were more knowledgeable and had better attitudes toward breast cancer screening than non-medical students [27].

These findings are plausible because students in health-related programs learn about breast cancer as part of their training. Therefore, more impactful programs on breast cancer awareness and screening should target students, especially those in non-health-related training programs.

### Strengths and limitations

This study used a previously validated questionnaire for the Zambia setting with objective knowledge and attitude measures. Additionally, the study leveraged the online platforms to reach students across the two main campuses of the University of Zambia thereby ensuring high response rate. However, this study has some limitations. This study was conducted at a single site, the University of Zambia, therefore these findings cannot be generalized to all the Universities. Additionally, the focus was on females’ student, and therefore future studies should involve men

## Conclusion

The participants’ overall level of knowledge of breast cancer screening was below average, highlighting a critical need for targeted educational interventions.

Additionally, the participants’ overall attitude toward breast cancer screening was moderate. This study revealed that the factors associated with good knowledge and attitude were a program of study.

If confirmed by larger studies, educational programs on breast cancer awareness and screening would mainly benefit University students in non-health-related training programs. Furthermore, deliberate efforts should be made to incorporate breast cancer screening programs and education in university programs.

## Data Availability

All relevant data are within the manuscript and its Supporting Information files

## Acknowledgements

The study team would like to acknowledge the contribution of the students and University administrators.

